# Dog-related deaths registered in England and Wales from 2001 to 2021

**DOI:** 10.1101/2022.10.14.22280913

**Authors:** John SP Tulloch, James A Oxley, Robert M Christley, Carri Westgarth

## Abstract

**Introduction:** This study aimed to describe the incidence and demographics of fatal dog bites or strikes, as defined in English and Welsh mortality data (2001-2021).

**Methods:** Data from the Office for National Statistics registered deaths dataset were used to identify individuals whose cause of death was defined as ‘bitten or struck by a dog’. The average annual number of dog-related deaths and trends in incidence were calculated. Age and sex demographics of victims were described.

**Results:** There were 3.3 (95%CI 0.3-6.3) dog-related deaths per year, a mean annual incidence of 0.59 (95%CI 0.06-1.11) deaths per 10 million population. There was no obvious trend in incidence. Of victims, 59% were male, 10% were <5 years, and 30% were ≥75 years.

**Discussion:** Dog-related deaths are rare in England and Wales and have not increased between 2001 and 2021. Further contextual information about the incidents is needed to be able to develop public health strategies and interventions.

**What is already known on this subject?:**

- Dog attacks can cause severe physical injury and psychological trauma, and can be fatal.
- Hospital admissions due to ‘dog bites or strikes’ in England have doubled between 1998 and 2018, with males and young children the most likely to be bitten.

**What this study adds?:**

- Deaths attributed to dog bites or strikes in England and Wales over a 20-year period are rare events and do not appear to be increasing in incidence.
- Registered deaths were more common in the very young and old, and in males.
- Further contextual information concerning dog-related deaths is needed to develop public health strategies and interventions.

## Introduction

Between January and 10^th^ October 2022 the media have reported that at least nine people have been killed by dogs in England and Wales, and have suggested that fatal attacks are increasing.[1] The incidence of dog-bite related hospital admissions in England more than doubled between 1998 and 2018, with over 8,000 individuals being admitted annually and males more likely than females.[2] For females, there were peaks in childhood (5-9 years) and in middle age (45-49 years), whilst for males, there was a sole peak in childhood (10–14 years). National data regarding emergency departments is not available but data from a tertiary paediatric hospital reported stable levels of attendance for a dog bite between 2016 and 2019, with no differences in attendance by sex; 33.4% of attendees were 7-12 years and 26.5% were 1-3 years.[3] It is unknown whether national dog-related deaths have increased in line with hospital admissions, and whether the demographics of the victims are similar.

This study aimed to describe English and Welsh mortality data where the cause of death was registered as a dog bite or strike. We hypothesised that mortality incidence trends would be rising similar to hospital admissions data.

## Methods

The Office for National Statistics (ONS) dataset ‘Deaths registered in England and Wales – 21^st^ century mortality’ was explored.[4, 5] These data are collated through the official certification and registration of deaths, a legal requirement since 1837, as such they have almost complete population coverage of deaths occurring in England and Wales.[6, 7] The data describe the annual number of registered deaths stratified by age group, sex, year, geographical region (only from 2013 onwards), and the underlying cause of death. The cause of death is decided upon by considering data from medical practitioners and coroners alongside information provided once a death has been registered. Cause of death is coded utilising the World Health Organisation’s International Classification of Diseases and Related Health Problems, Tenth Revision (ICD-10).[8]

Deaths were identified using the ICD-10 code group W54 (Bitten or struck by a dog). This was the same code group previously used to identify individuals admitted to English hospitals due to dog bites.[2] The annual average number and population incidence of dog-related deaths were calculated using ONS population data as the denominator,[9] and trends were described. Incidence was stratified by geographical region and plotted. The age and sex of the victims were described and differences in sex-stratified incidence were examined using χ^2^ analysis. The ICD-10 codes used were analysed to identify any contextual information about the location of where the dog bites occurred (Table 1).

**Table 1.** Frequency of ICD-10 codes used to register dog-related deaths in England and Wales 2001-2021 (* This was the sole code used since 2020).

| ICD-10 code | Description | Number |
| --- | --- | --- |
| **W54\*** | Bitten or struck by dog | 8 |
| **W54.0** | Bitten or struck by dog: home | 34 |
| **W54.1** | Bitten or struck by dog: residential institution | 0 |
| **W54.2** | Bitten or struck by dog: school, other institution and public administrative area | 0 |
| **W54.3** | Bitten or struck by dog: sports and athletics area | 0 |
| **W54.4** | Bitten or struck by dog: street and highway | 8 |
| **W54.5** | Bitten or struck by dog: trade and service area | 0 |
| **W54.6** | Bitten or struck by dog: Industrial and construction area | 0 |
| **W54.7** | Bitten or struck by dog: farm | 0 |
| **W54.8** | Bitten or struck by dog: other specified places | 6 |
| **W54.9** | Bitten or struck by dog: unspecified place | 13 |
| **Total** |  | **69** |

All statistical and spatial analyses were carried out using R language (version 3.2.0) (R Core Team 2015). Results were deemed statistically significant where p < 0.05. No ethical approval was needed as this was an analysis of publicly available data, with no personally identifiable information. Patients were not involved in the design, nor participated in the research, of this study.

## Results

Between 2001 and 2021, 69 individual deaths were registered as being caused by a dog bite or strike. The annual number of deaths were approximately normally distributed, the mean number of dog-related deaths per year was 3.29 (95%CI 0.30-6.27).

The mean annual incidence was 0.59 (95%CI 0.06-1.11) per 10 million (Fig 1). The highest incidence of dog-related deaths was in 2011, with 1.07 (95%CI 0.44-2.20) per 10 million population. There was no obvious trend in incidence.

**Figure 1.**
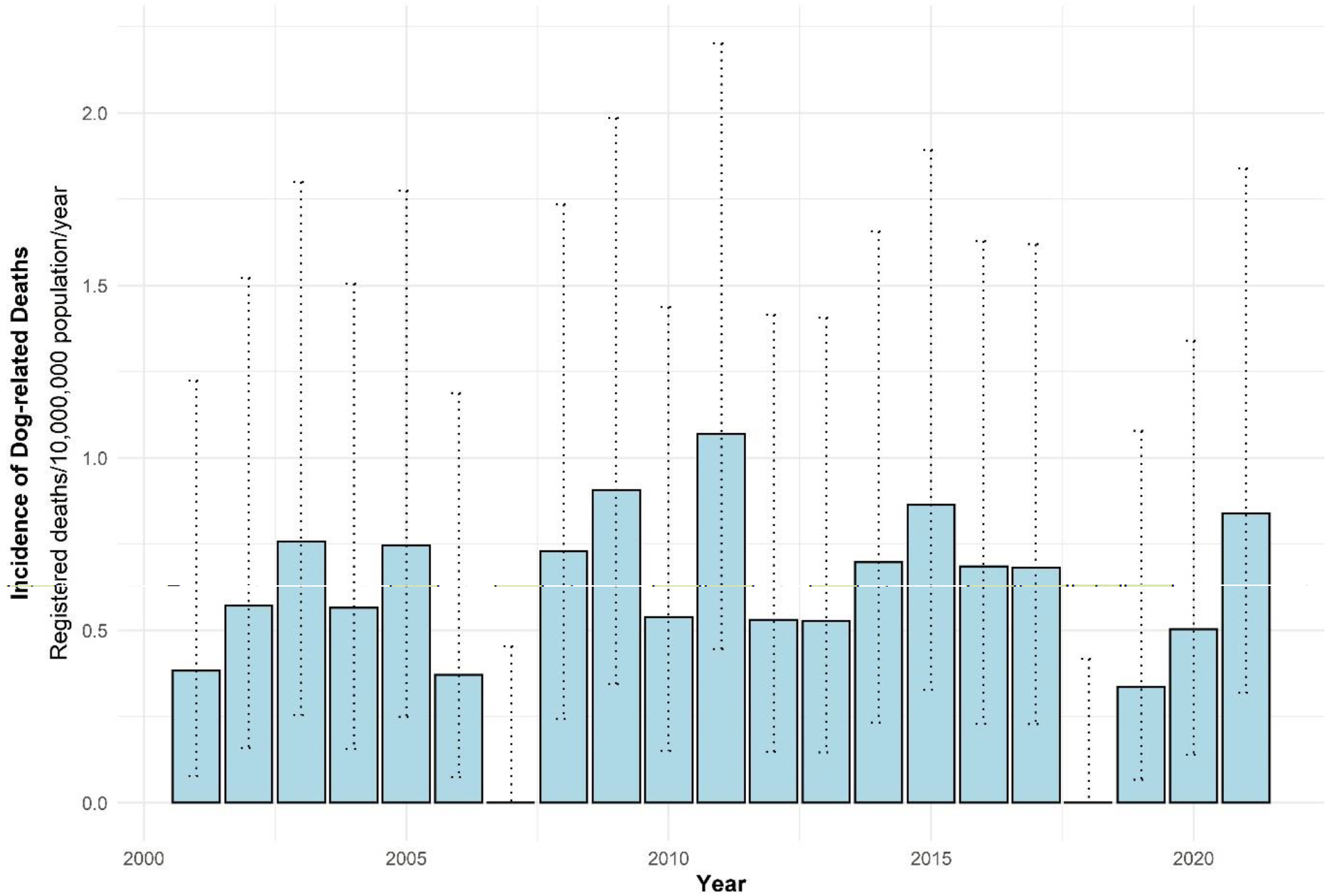
The annual incidence (death per 10 million population) of dog-related deaths in England and Wales (2001-2021). Dashed lines represent 95% confidence intervals.

Regional variation of dog-related deaths incidence was evident. The North-West of England had the highest average annual incidence of 1.36 deaths per 10 million population (95%CI 0.67-2.48), and the East of England had the lowest, 0.18 deaths per 10 million population (95%CI 0.02-0.83) (Fig 2).

**Figure 2.**
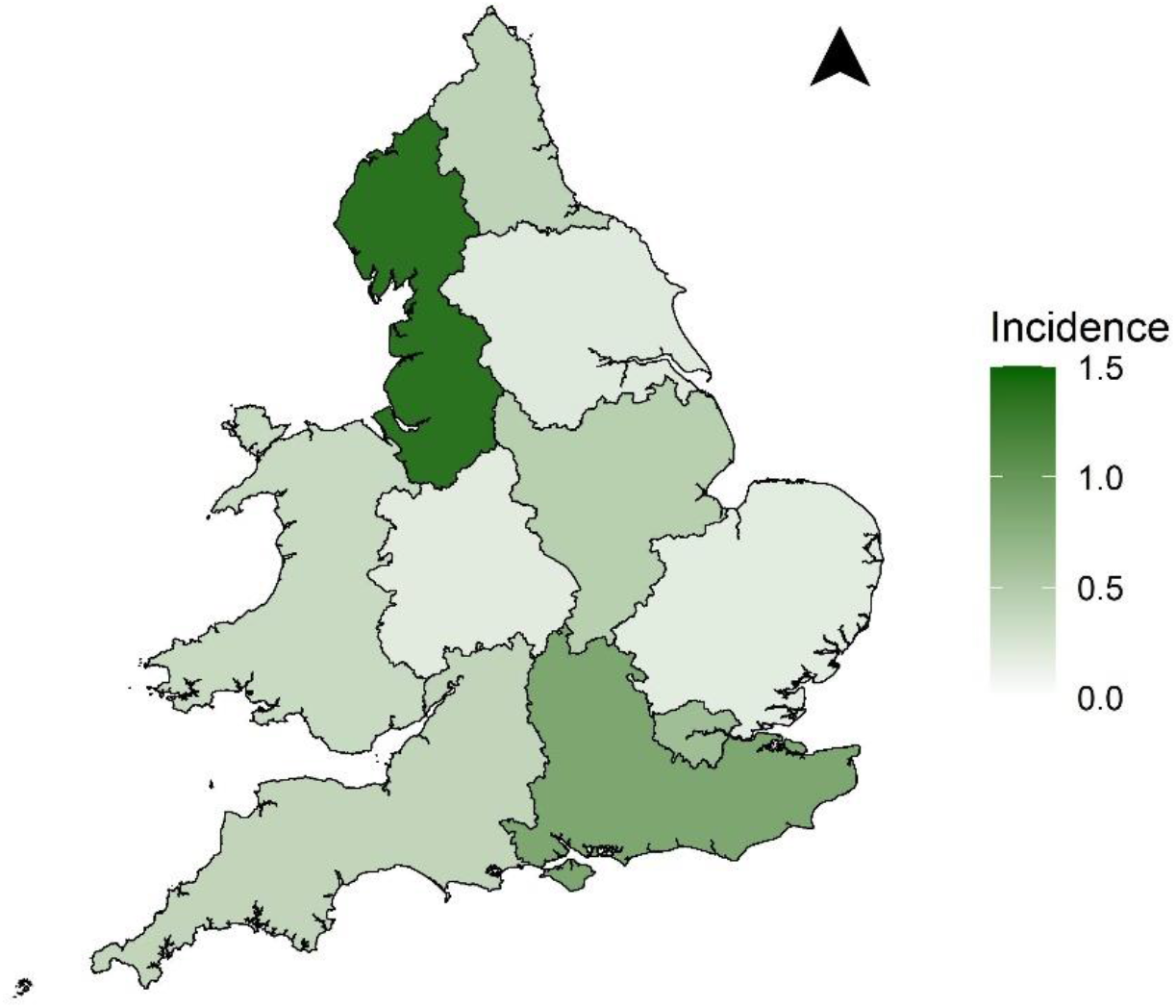
Average annual incidence (cases per 10 million population per year) of dog-related deaths in England and Wales (2013-2021).

Fifty-nine percent of victims (n=41, 59.4% 95%CI 46.9-71.1) were male (0.67 cases per 10 million population (95%CI 0.48-0.91), compared to 0.44 cases per 10 million (95%CI 0.29-0.63) in females (p=0.08)). There appeared to be three peaks in incidence among males; 75-84 years (75-79 = 2.01, 80-84 = 2.36), 50-54 years (1.02), and <5 years (0.86) (Fig 3) and two in females; 75-79 years (1.53), and <5 years (1.21).

**Figure 3.**
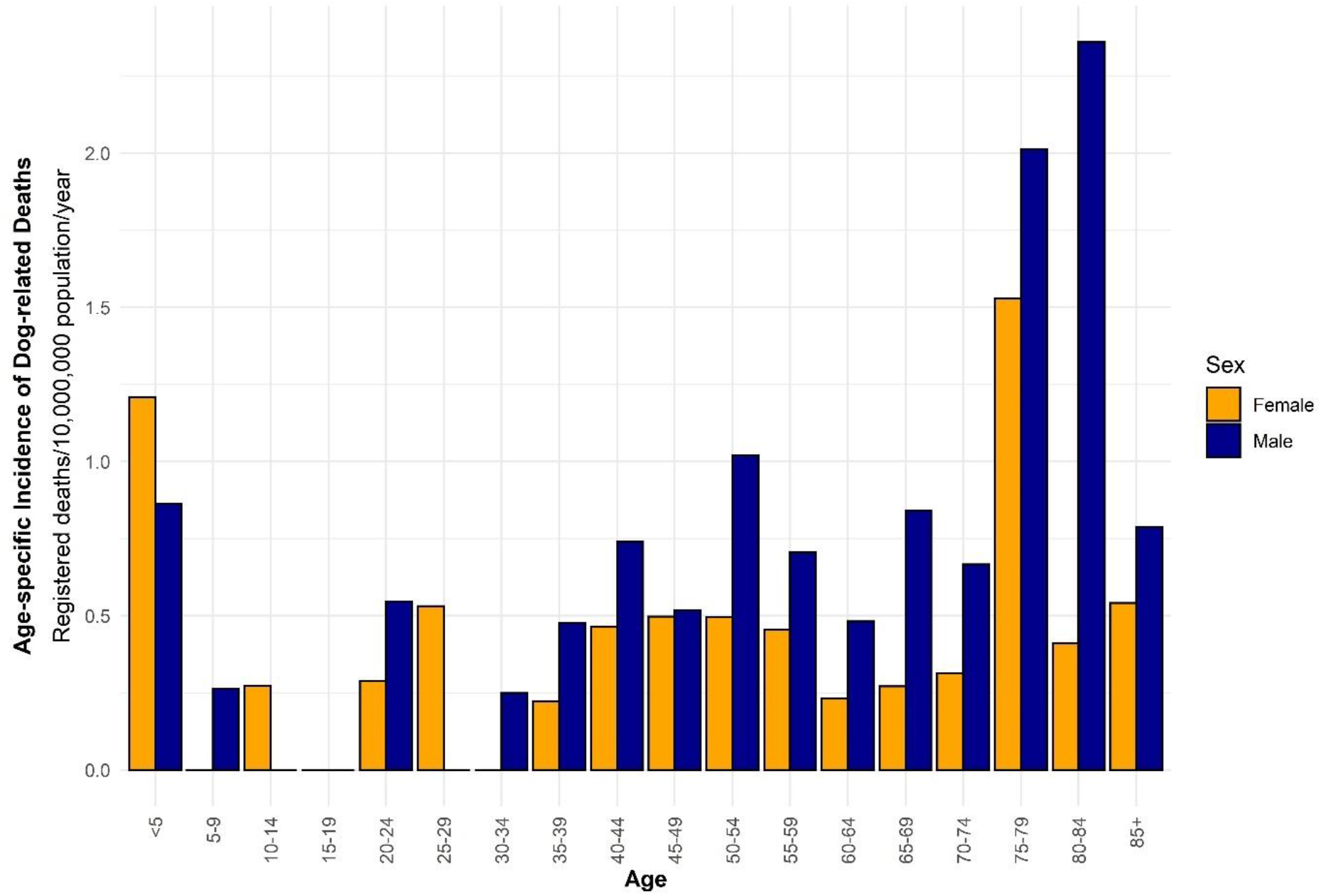
Annual age-specific incidence (deaths per 10 million population) of dog related deaths in England and Wales stratified by sex (2001-2021).

The majority of ICD-10 codes were not used, and from 2020 onwards the summary code ‘W54’ was solely used (Table 1). Only two codes specified a location of the dog attack (n=42, 60.9%). Of these, 81.0% (95%CI 65.9-91.4) occurred at home and 19.0% (95%CI 8.6-34.1) occurred on the street or highway.

## Discussion

The findings of this study do not support our initial hypothesis that dog-related deaths in England and Wales were increasing in number between and including 2001-2021, and instead they appear to be at a stable, low rate; averaging three deaths per year. For context, over the same period on average one person per year died due to a lightning strike (ICD-10 code: X33),[4] 3 after falling from a tree (ICD-10 code: W14), [4] and 1,548 in road accidents.[10] Data has yet to be officially collated for 2022 but it may record the greatest number of deaths for over 20 years. However, this must be treated with caution as one anomalous year does not make an increasing trend. Nevertheless, the media reports are concerning, and we recommend that data need to be reviewed annually to identify whether any trends of dog-related deaths emerge.

The calculated incidence of dog-related deaths (0.59 deaths per 10 million population) is slightly lower than in the United States of America (USA) (1.1 deaths per 10 million population),[11] and similarly stable.[12] However, the annual European Union (EU) incidence, between 1995 and 2016, has been estimated as 0.9 per 10 million population, with an annual increase of 5.6%,[13] ranging from 0 in Ireland and Luxembourg to 4.5 in Hungary, with a median of 0.6 deaths per 10 million across the EU nations. It can be concluded that the incidence of dog-related deaths in England and Wales are not anomalous and is similar to many other North American and European countries. However, the trend data do not match the EU, which saw an increase above and beyond the growth in the dog population.[13] This is similar to English hospital admissions for dog bites, which saw an increasing rate of admissions that was greater than rate of growth in the dog population.[2]

Furthermore, across many North American and European nations, victims are predominantly male, with peak deaths in young children and the elderly.[12–15] Numerically more males were seen here, but this was not statistically significant. Here there was a high proportion of deaths seen in the <5 years (10.1%) and ≥75 years (30.4%).

There appears to be a disconnect between the stable incidence of dog-related deaths and the increasing incidence of hospital admissions for dog bites in England, which more than doubled over a similar period.[2] It is unknown why dog-related deaths are not increasing at the same rate. It may be that dog bites leading to minor injuries are increasing, but the most severe injuries (including fatal ones) are not increasing. Without further contextual and medical information about the nature of the bite or attack and the type and severity of injury, it is not possible to draw a conclusion.

The geographical spread of deaths were inconsistent with the spatial distribution of hospital admissions.[2] Hospital admissions data identified hotspots in areas that would have equated to the regions of the North West (similarly highest death incidence out of the 10 regions), and North East (only 5^th^ out of the 10 regions), whilst the area with the lowest hospital admissions was London (3^rd^ highest death incidence). This is most likely due to the low number of deaths in each region resulting in the modifiable area unit problem form of bias. As such, these spatial patterns must be considered with a degree of caution.

The demographics of those killed versus those admitted to hospital also differed slightly. In both cases, peaks were seen in young children, but were younger in the registered deaths dataset.[2] Peak adult admissions occurred in middle age (40-49 years), whilst peak adult deaths occurred in >75 years. This is likely due to the increased vulnerability of younger children and the elderly to fatal injury. The type of location of the dog-related incident, as recorded by ICD-10 codes, is comparable with 81.0% of individuals killed at home compared to 83.9% of those admitted to hospital, and 19% and 12% on the street, respectively.[2]

The major limitation of these data is the lack of contextual information about the events leading up to the victims’ death. Understandably, as this is not the dataset’s primary purpose, there is no data concerning the dogs involved, the nature of the attack and events preceding it, and the extent and severity of injury. From a public health perspective, these data are critical so that interventions can be developed that may prevent these rare events from happening in the future. Due to the low number of deaths, when these data are stratified by demographics or geographical areas, group size becomes small and so only crude descriptive analysis can occur.

## Conclusions

Dog-related deaths are very rare events within England and Wales and have not been increasing in incidence to 2021. The incidence and demographics are broadly similar to those of other European and North American nations. More contextual information is needed about dog-related deaths to be able to elicit change and develop effective public health strategies.

## Data Availability

All data produced are available online at https://www.ons.gov.uk/peoplepopulationandcommunity/birthsdeathsandmarriages/deaths/datasets/the21stcenturymortalityfilesdeathsdataset

## Contributions

JT conceived and designed the work, analysed the data, and drafted the manuscript. All authors interpreted the data, helped to revise the manuscript, and approved the final submitted manuscript. All authors agree to be accountable for the accuracy and integrity of this work.

